# Monitoring the post-containment COVID-19 crisis in Guadeloupe: Early-warning signals of destabilisation through bootstrapped probability density functions

**DOI:** 10.1101/2020.05.29.20117333

**Authors:** Meriem Allali, Patrick Portecop, Michel Carlès, Dominique Gibert

**Affiliations:** Emergency Medical Service (SAMU-SAU), University Hospital, Pointe à Pitre, Guadeloupe; Critical Care Department, University Hospital, Pointe à Pitre, Guadeloupe; Univ Lyon, Univ Lyon 1, ENSL, CNRS, UMS 3721, LGL-TPE, F-69622 Villeurbanne, France

**Keywords:** COVID-19, Monitoring, Guadeloupe, SAMU, Monte Carlo Model, Spread control, Quarantine, Early warning, Bifurcation, Bootstrap, Probability density functions, Outlier, Change point model

## Abstract

We propose a method to detect early-warning information in relation with subtle changes occurring in the trend of evolution in data time series of the COVID-19 epidemic spread (e.g. daily new cases). The method is simple and easy to implement on laptop computers. It is designed to be able to provide reliable results even with very small amounts of data (i.e. ≈ 10 − 20). The results are given as compact graphics easy to interpret. The data are separated into two subsets: the old data used as control points to statistically define a “trend” and the recent data that are tested to evaluate their conformity with this trend. The trend is characterised by bootstrapping in order to obtain probability density functions of the expected misfit of each data point. The probability densities are used to compute distance matrices where data clusters and outliers are easily visually recognised. In addition to be able to detect very subtle changes in trend, the method is also able to detect outliers. A simulated case is analysed where *R*_0_ is slowly augmented (i.e. from 1.5 to 2.0 in 20 days) to pass from a stable damped control of the epidemic spread to an exponentially diverging situation. The method is able to give an early warning signal as soon as the very beginning of the *R*_0_ variation. Application to the data of Guadeloupe shows that a small destabilising event occurred in the data near April 30, 2020. This may be due to an increase of *R*_0_ ≈ 0.7 around April 13–15, 2020.

## Introduction

By mid-May 2020, the COVID-19 disease caused by SARSCoV2 continues its worldwide spreading with nearby 4 millions of people infected and 300,000 deceased persons (1). Faced with this rapid and hardly controllable epidemic disease, many countries decided to close they frontiers, and to strongly reduce their industrial and economic activities to decrease as much as possible the contagiousness of their population. Although these measures may take different forms, many countries adopted the containment solution, i.e. the lock down of people at home, to quickly and strongly enhance social distancing in order to reduce the transmission of the virus among people (2, 3). Such is the case of France, where the containment started on March 17, 2020 and is scheduled to be partly released on May 11, 2020, i.e. a duration of 55 days. On the overall, the containment proved efficient, with a net decrease of the number of patients in critical situation and necessitating intensive cares. This enabled to avoid a collapse of the sanitary facilities, particularly the intensive care units. The efficiency of the containment is variable from one region to another, depending on the sanitary situation at the beginning of the containment. As shown in our previous study (4), the case of Guadeloupe was particularly favourable to a good control of the epidemic spread due to several factors: i) the small number of infected persons at the beginning of the containment, ii) a good and coherent communication by the sanitary and administrative authorities, iii) a good respect of the social distancing rules by a vast majority of the population, iv) a tight control of the incoming passengers by either airport or ship.

Depending on the number of supposed infected people at the end of the containment period, the sanitary procedures to apply are different. Such is the case in France, where the eastern regions, including Ile-de-France around Paris, are tagged as “red” (i.e. with a widespread circulation of the virus among the population) while all other regions, excepted Mayotte, are declared “green”. In Allali et al., we show that the situation in Guadeloupe at the beginning of the post-containment period will correspond to a pre-epidemic state with a small number of infected people and an essentially non-infected population forming a large reservoir of “susceptible” persons (ie who may be infected). Since no vaccine is yet available and because of the high vulnerability of the population, the post-containment period is particularly critical and a restart of the epidemic spread, the so-called second wave, is inevitable if no social distancing and no sanitary control is performed (6). In Allali et al. (5), we tested a variety of solutions combining social distancing and systematic placement of symptomatic patients in quarantine. We also considered the detection and placement in quarantine of asymptomatic persons by means of guided testing in the entourage of the detected symptomatic people. As discussed by Allali et al. (5), the model is mainly constrained by three key parameters: *R*_0_, *N_a_* and *δT_Q_*. Here, *R*_0_ is the basic reproduction number which may be decomposed into several components (see (5) for a detailed discussion). *N_a_* is the average number of asymptomatic persons that are supposed to be detected and placed in quarantine each time one symptomatic patient is identified. *δT_Q_* is the time delay necessary to isolate a new symptomatic patient together with the asymptomatic identified in her/his entourage. A main result of our study (5) is that, depending on the values assigned to the *R*_0_, *N_a_* and *δT_Q_* parameters, the spread control may be either stable or unstable. This allowed us to identify sub-domains in the 3D solution space with coordinates (*R*_0_*,N_a_,δT_Q_* (Fig. 1). In the case where a stable solution is obtained, the exponential epidemic spread is rapidly stopped and followed by a sharp damping with a small number of patients needing intensive cares. An example of such a damping is shown in Figure 2B for the Turkish data. In the case of an unstable solution, the initial exponential increase is not damped and the dreaded second wave occurs. We also show that the stable and unstable solution domains are separated by a narrow critical boundary where the effective basic reproduction number *R*_0_ ≈ 1 (1). On this boundary, both the exponential divergence and the sharp damping are absent, and they are replaced by oscillating characteristics (i.e. number of new infected, critical cases and deceased persons) with a period of one or two weeks. Interestingly, this seems to be case of the situation both in the United States of America (Fig. 2A) and in Sweden (Fig. 2B) where strong oscillations are observed in the time series of the number of daily new deaths. The reader can compare these time-series with the synthetic curves of Figure 6A-D in Allali et al. (5). The oscillations visible in the curves for United States of America and for Sweden may not be attributed to seasonal effects (7) but could be due to stochastic resonance (8). Such a critical situation should be avoided because the absence of damping produces an ever-growing number of deceased persons. Ultimately, the entire population will have been contaminated.

**Fig. 1.**
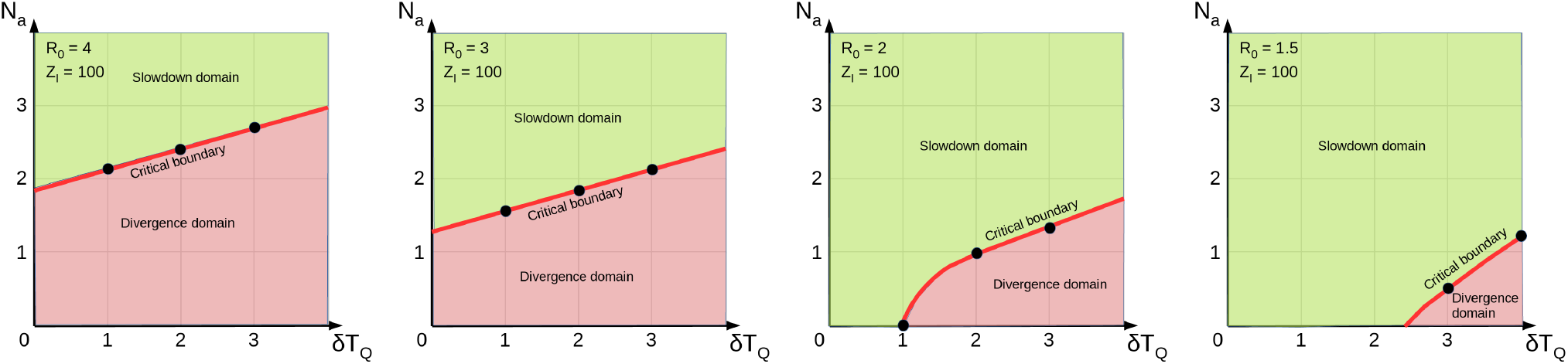
Solution domains (*δT_q_,N_a_*) obtained for *R*_0_ = 4,3,2,1.5. *δT_Q_* is the time-delay elapsed between the apparition of symptoms in a patient and the moment when this patient is placed in quarantine. *N_a_* is the average number of asymptomatic persons detected and placed in quarantine when a symptomatic patient is detected. The green domains represent the models for which the epidemic spread is controlled. The red domains are the set of model for which the epidemic spread is uncontrolled with an exponential growing. The solid red lines represent the boundary between both domains and where the models are unstable with oscillations and large fluctuations.

**Fig. 2.**
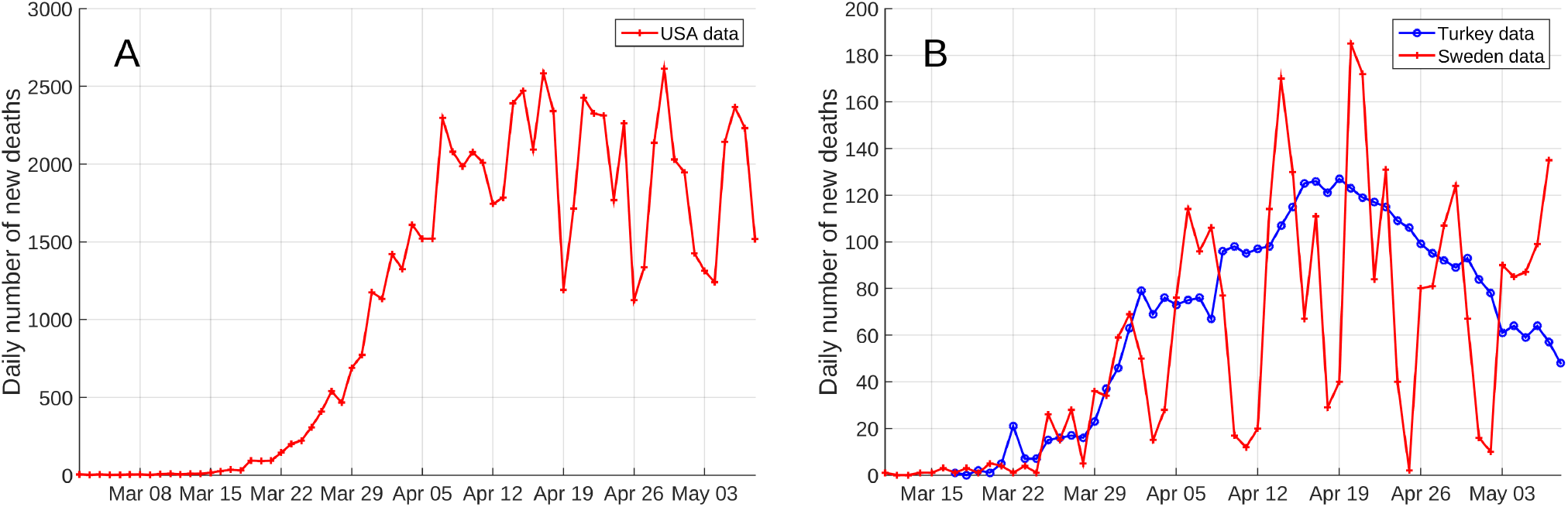
Daily number of new deaths recorded in USA (A), and in Turkey and Sweden (B). The conspicuous oscillations in the USA and Sweden curves are typical of a critical equilibrium near a tipping point (i.e. somewhere on a critical boundary in Fig. 1. The bell-shaped curve of Turkey is typical of a stable damping of the epidemic spread (i.e. somewhere in a green domain in Fig. 1). Data are from (9).

In the present study, we address the problem of the real-time monitoring of the epidemic situation during the post-containment period. The aim of this monitoring is to control that the situation remains in the domain of stable solutions as described by Allali et al. (5) and that it does not evolves dangerously toward the critical boundary. In the next Section, we present some simulations performed with the stochastic model developed in Allali et al. (4). These simulations are performed with a time-varying *R*_0_ in order to cross the critical boundary from a stable solution to an unstable one. Then, we propose a detection method able to detect subtle changes in the data time-series and susceptible to constitute early-warning signals indicating that the sanitary situation is evolving toward an unstable configuration. We develop this method by keeping in mind that it must be as simple as possible and easy to implement.

## Method

This Section presents the method used hereafter and designed to help detecting early-warning signals of possible destabilisation of the COVID-19 spread control during the post-containment period.

The aim of the method is to help to detect as early as possible subtle changes in the time-series of available data (i.e. daily or cumulative number of new infected, new deaths, etc.). There exist a huge amount of methods able to detect subtle changes, invisible for the human eyes, in time series (see e.g. (10, 11) for synthetic and real data examples). However, most of these methods are not suitable for our purpose, mainly because we have to deal with a very small number of data points (see e.g. Royall (12) for a discussed on the pitfalls of small-sample statistics). This is the main reason why we developed a purposely designed method able to tackle with this particularity. The method discussed hereafter may constitute the algorithmic core of change-point detection, outlier signalling and clustering of data sets.

The problem we address is explained in Figure 3. Suppose that we have a small number of data points {*d*_1_*,d*_2_*,…,d_N_*} forming a short time-series corresponding, for instance, to the number of new infected people. Using these points, we want to check if the most recent data points, hereafter called the “test” points (red dots Fig. 3), significantly depart from the trend (purple line Fig. 3) defined by the older data points, called the “control” points (black dots Fig. 3). We admit that the older data used to determine what we call the trend may contain some outliers that we also want to identify.

**Fig. 3.**
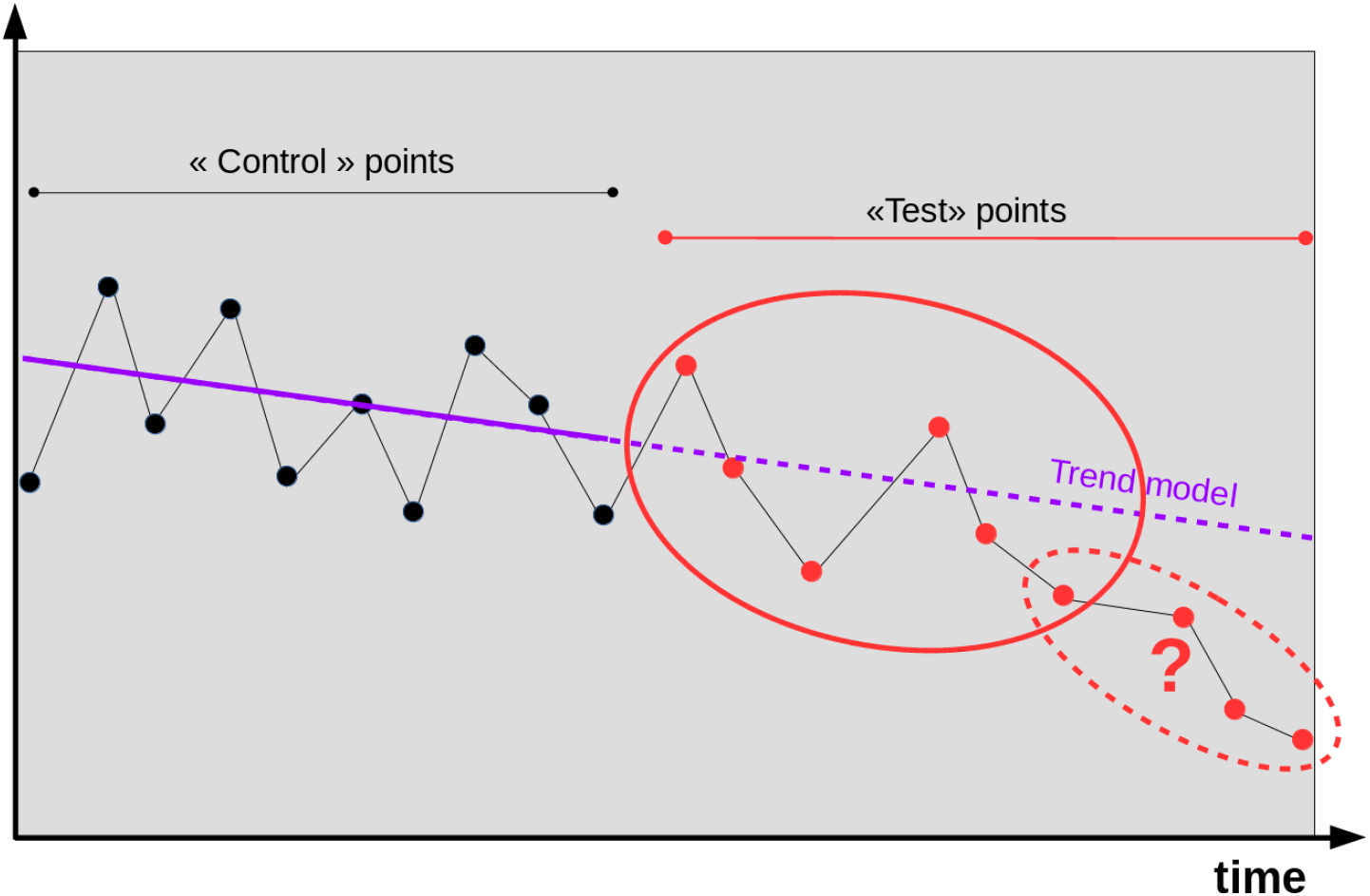
Principle of the method proposed in the present study. A small data set is separated into two subsets. The subset of “control” points (black dots) is used to define a trend (solid straight line in purple). The trend is continued (dashed purple line) toward the subset of “test” points (red dots) in order to test whether or not these “test” points remain coherent with the trend of the “control” points. The method should be able to distinguish coherent “test” points (in the elliptic solid line) from incoherent “test” points (elliptic dashed line). The method should also be able to determine the change-point (marked with a red ?) as precisely as possible.

As stated above, we want to design an as simple method as possible in order to give the user the maximum allowance of adaptation to the data at hand depending on her/his expertise. As illustrated in Figure 3, the control points are scattered and, depending on the importance (the weight) given to each control point, the definition of the trend is fuzzy. The idea at the root of the method is to explore the set of all acceptable trends by using a bootstrap technique where a very large number of trends are determined by assigning different weights 0 ≤ *w_i_* ≤ 1 to the control points (Fig. 4A). The misfits *δ_i_* between the trends and the data points *d_i_* (Fig. 4B) are stored and subsequently used to compute the normalised “discrepancy histograms” hereafter called “probability density functions” (pdf’s) *f_i_*. The pdf’s are used to statistically identify clusters of coherent and incoherent data points (Fig. 4C).

**Fig. 4.**
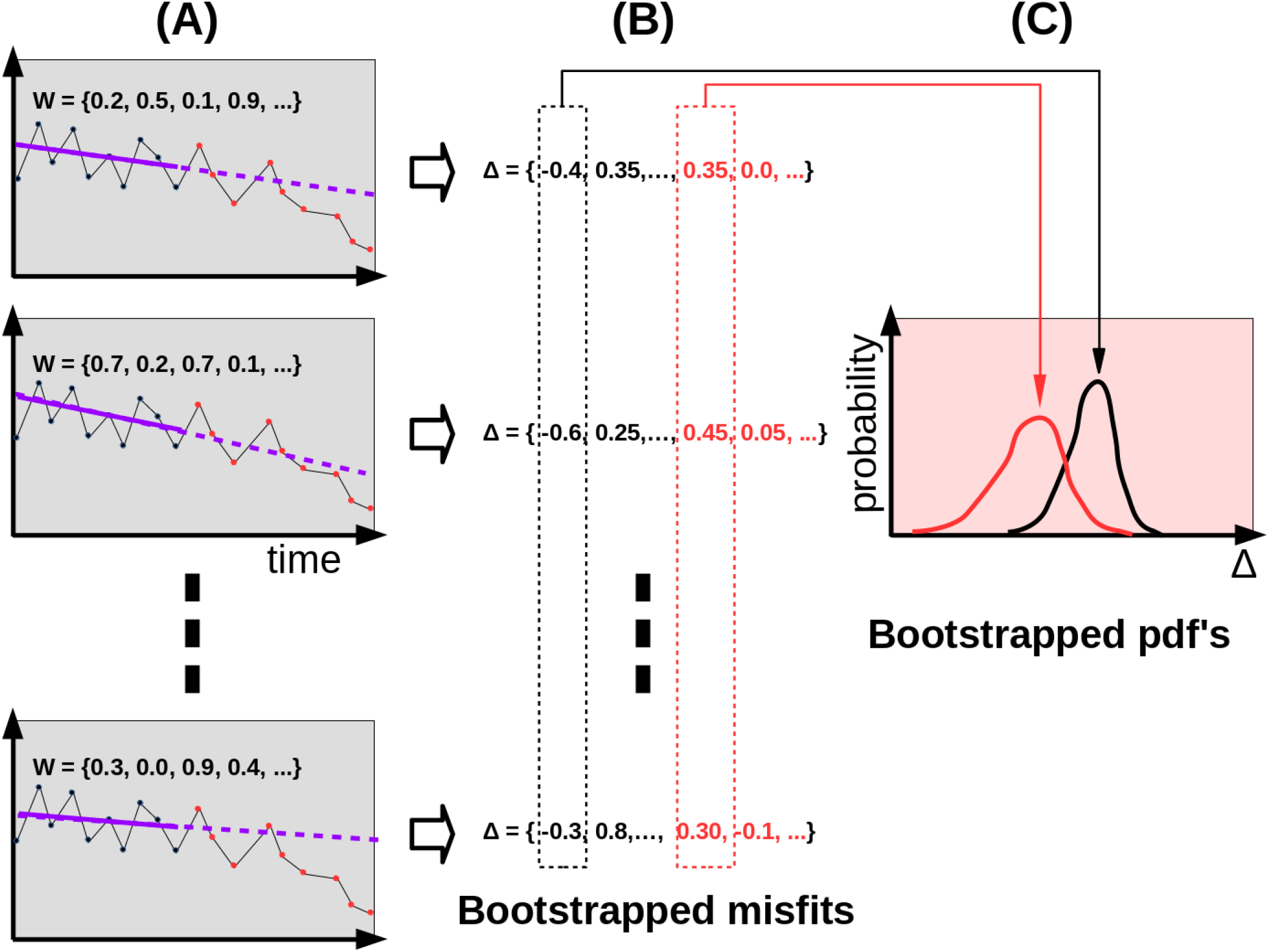
Principle of the bootstrap implemented in the method. A) The trend model is fitted to the control points a great number of times with different random weights 0 ≤ *w_i_* ≤ 1 assigned to the control points. B) The bootstrapped misfits (i.e. the discrepancies between the data points and the trend) are stored. C) The misfits are used to construct a normalised histogram (i.e. a probability density function) for each data point.

Figure 5 shows the results for the different stages of the computational procedure. In this example, the experimental recovery of the probability density functions is achieved by fitting a trend model to the subset of control data points (here points number 1 to 9) including the outlier *d*_6_. Bootstrap is done by fitting the trend model a very large number of times (≈1000 times) with real random weights assigned to the data points (i.e. stage A of Fig. 4). The choice of the trend model is free and depends on the particular data set to analyse. In the present example, we use polynomials of degree one (i.e. straight lines) and a least-squares fit. This choice may be changed according to the data at hand. Figure 5B,C shows two such fits. Because of the different values given to the weights, the best-fit lines differ from one fit to the other (blue solid lines). It is the same for the misfits represented by the vertical dashed segments connecting each data point to the best-fit line.

**Fig. 5.**
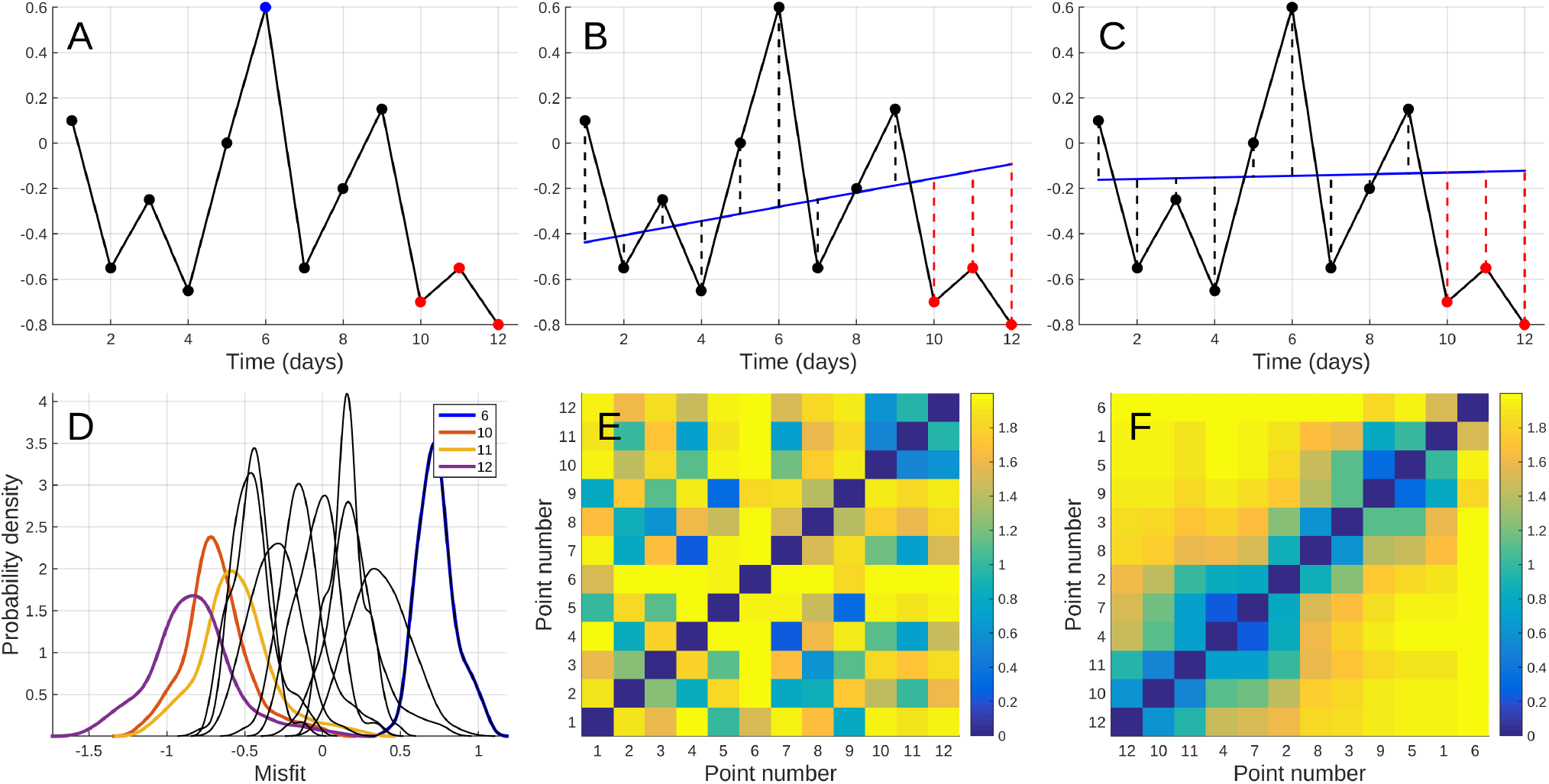
Implementation of the method proposed in the present study. A) A small set of data points is separated into two subsets: the “control” points (black dots) and the “test” points (red dots). In the case of on-line change-point detection, the “test” points are the more recent data and the “control” points are the oldest data. B) The “control” points are used to define a trend model, here a straight line (blue line) adjusted by least-squares fit. The adjustment of the trend is done by giving a random weight 0 ≤ *w* ≤ 1 to each “control” point (e.g. a data point with *w* = 0 is not considered in the adjustment). The discrepancies between the fitted line and the data points (vertical dashed lines) are stored to subsequently construct the probability density functions (pdf’s) shown in (D). C) Another example of trend adjustment obtained with different weights assigned to the data points: the discrepancies are different from those obtained for case (B) and they are stored to compute the pdf’s. D) Experimental probability density functions *f_i_* obtained by normalising the histograms of the discrepancies obtained for each data point. E) Matrix **D** filled with the distance between each pairs of pdf’s. Pixel (*i,j*) corresponds to the distance *L*_1_(*f_i_,f_j_*) (eq. 3) between pdf *f_i_* and pdf *f_j_*. The diagonal is filled with zeros because *L*_1_(*f_i_,f_i_*) = 0. For example, column 9 and line 9 of the matrix give the distances between pdf *f*_9_ and all pdf *f*_1_*,f*_2_*,…,f*_12_. The colour bar goes from 0 (blue) to 2 (yellow). F) Reorganised matrix **D**\* obtained by permuting the lines and the columns of **D** so that the entropy (i.e. complexity) of **D**\* is minimum. This reorganisation may (but not systematically) be useful to enhance particular patterns that are not apparent in the original **D** matrix. Graphs B and C correspond to stage A in Figure 4, and graph D corresponds to stage C in Figure 4.

The collection of misfits obtained during the bootstrap iterations (i.e. stage B of Fig. 4) are subsequently used to construct normalised histograms (i.e. stage C of Fig. 4) which approximate the probability density functions *f*_1_*,f*_2_*,…,f*_12_ (Fig. 5D). At this stage of the processing chain, some interpretation is already made possible. For instance, it can be observed that the pdf of data 6 (blue curve on the right part of the plot) seems to depart from the set of pdf corresponding to the other points used to perform the fits (black curves grouped toward the centre of the graph). A quantitative assessment of the discrepancies between pdf *f_i_* and *f_j_* may be obtained with measures like, for instance, the Kullback-Leibler information (see e.g. Section A1–4 of Martinez et al. (13)),

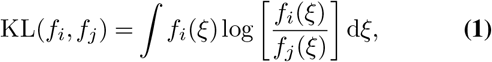

the Jenssen-Shannon divergence (14), also called the information radius,

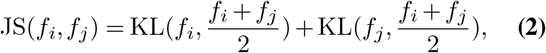

and the *L*_1_ norm,

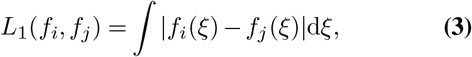

which is twice the total variation distance.

The KL measure is non-symmetrical and may take infinite values. It is not a metric and does not satisfy the triangle inequality. Such is not the case of JS which is symmetrical and is a metric such that JS = 0 for identical distributions and JS = 2log2 for maximally different distributions. The *L*_1_ norm is well-defined for arbitrary probability density functions and is such that 0 ≤ *L*_1_(*f_i_,f_j_*) ≤ 2, the minimum value corresponds to *f_i_* = *f_j_* and the maximum value is obtained when the supports of *f_i_* and *f_j_* are disjoint.

The distance measures defined in equations 1–3 may be used to compute a symmetric distance matrix **D** filled with the distances of all pairs (*f_i_,f_j_*) that can be formed with the set of experimental pdf like those shown in Figure 5D. In the present example, the matrix **D** is computed with the *L*_1_ norm and counts 12^2^ entries, including the diagonal self-distances (i.e. *L*_1_(*f_i_,f_i_*) = 0 for *i* = 1,…,12).

Figure 5E shows the **D** matrix represented in its natural time ordering, i.e. with the lines and columns ordered according to the time index. The distance matrix only contains a part of the information brought by the pdf’s of Figure 5D, however this representation allows to easily identify clusters of coherent data points that are grouped in a time interval. A quick look at Figure 5E reveals the existence of a small cluster of short distances that appears as a blue square in the upper-right part of the matrix. This cluster corresponds to the data points {*d*_10_*,d*_11_*,d*_12_} which are statistically coherent and grouped on the time axis. The remaining parts of the matrix are less structured, with pixels of small to high distances (blue to yellow).

More patterns can sometimes be seen in the matrix after reordering the columns and the lines in order to minimise the entropy of the image. This is simply achieved by permuting the lines and the columns in the same way in order to keep the matrix structure. The rearranged matrix **D**\* is represented in Figure 5F. We recognise the cluster of data points {*d*_10_*,d*_11_*,d*_12_} in the lower-left corner of the matrix (this particular position is not mandatory). However, other features now appear, with a diagonal band of low-distance pixels showing that the “control” data points {*d*_1_*,…,d*_9_} used to fit the trend model during the bootstrap are statistically coherent. A noticeable exception is point *d*_6_ which locates in the upper right corner of **D**\* and shares no low-distance pixel with any other data point (i.e. both the line 6 and the column 6 of the matrix do not contain blue pixels). We may then conclude that data point *d*_6_ is an outlier. The fact that the cluster of blue pixels {*d*_10_*,d*_11_*,d*_12_} is not clearly disjoint from the blue diagonal band indicates that some data points in {*d*_10_*,d*_11_*,d*_12_} are not all statistically disjoint from some points in {*d*_1_*,d*_2_*,d*_3_*,d*_4_*,d*_5_*,d*_7_*,d*_8_*,d*_9_}. Indeed, a careful examination of the lower-left part of **D**\* reveals that data {*d*_10_*,d*_11_} share blue/green pixels with data {*d*_4_*,d*_7_}. Such is not the case of point *d*_12_ which only shares blue pixels with its companions of cluster, i.e. the data points {*d*_10_*,d*_11_}.

To conclude this methodological section, we may claim that our detection method is able to reliably identify both outliers and data points marking a change with respect to a trend defined by older data. This identification relies on an examination of distance matrix represented in its natural ordering (Fig. 5E) and in its low-entropy reordered configuration (Fig. 5F). Thanks to the bootstrap, it is possible to process very small data sets, a characteristic mandatory to perform an online analysis of the data.

## Analysis of a simulated destabilisation of spread control

We now address the main question of the present study, namely the online analysis of a simulated destabilisation of the COVID-19 spread control. We use the stochastic computer code developed in (4, 5) to simulate a situation where we start from a stable convergent solution to cross the critical boundary in the solution domain (Fig. 1. As recalled in the Introduction, and discussed in details by Allali et al. (5), the critical boundary may be crossed by varying a single or several of the *R*_0_, *N_a_* and *δT_Q_* parameters. In the simulation discussed in the present study, we chose to vary *R*_0_ while keeping both *N_a_* and *δT_Q_* constant. The choice to vary only *R*_0_ is motivated by the desire to simulate a likely situation where social distancing is gradually deteriorates inside the population of Guadeloupe. That may be due to some overestimate of the safety of the situation which incites people to be less vigilant. Such an amplification of the epidemic risk is comparable to the well-known fact that accidents and catastrophes often occur at the end of a dangerous period, when people abusively consider they are now safe.

We define a trajectory starting at point *P_S_* = (*R*_0_ = 1.5*,N_a_* = 1*,δT_Q_* = 3) corresponding to a stable and damped control of the epidemic spread. Point *P_S_* is located in the green domain of the *R*_0_ = 1.5 graph of Figure 1. The end point of the trajectory *P_E_* = (*R*_0_ = 2.0*,N_a_* = 1*,δT_Q_* = 3) is located in the unstable red domain in the *R*_0_ = 2.0 graph of Figure 1. In order to simulate a situation where an insidious and subtle change of stability conditions occur, we linearly change *R*_0_ from 1.5 to 2.0 between day 31 and day 50 of the simulation. The values of the other parameters are given in Table 1.

**Table 1.** Model input parameters. The three first top parameters in the Table (*R*_0_, *δT_Q_, N_a_*) have changing values in the different simulations. Other parameters remain unchanged for all simulations.

| Parameter name | Symbol | Value | Comment |
| --- | --- | --- | --- |
| Basic reproduction number | $R_0$ | $1.5, \dots, 2$ | Time-varying parameter through the contagiousness function $\xi_c$. |
| Detection delay | $\delta T_Q$ | 1, 2, 3, 4 | This parameter is a time difference defined as the delay (in days) between the apparition of symptoms in a patient and the moment when this patient is placed in quarantine. This represents the time delay taken by the sanitary authorities to tackle stop the contagiousness of a symptomatic patient. |
| Number of detected asymptomatic | $N_a$ | 0 to 4 | Number of asymptomatic people detected in the entourage of each detected symptomatic patient. |
| Number of initial asymptomatic | $Z_I$ | 1000 | |
| Switching function (lognormal) | $\zeta_{a/s}$ | $\mu_\zeta = 5.6$ (days)<br>$\sigma_\zeta = 3.9$ (days) | This probability function give the switching time dependence of the contagiousness of asymptomatic, pre-symptomatic and symptomatic people. $\zeta_{a/s}$ is taken as a lognormal distribution with mean and standard deviation given by Ferretti et al. (17). |
| Contagiousness function (gamma) | $\xi_c$ | $\alpha_\xi = 5$ (shape)<br>$\beta_\xi = 1$ (scale) | Represents the time dependence of the contagiousness of asymptomatic, pre-symptomatic and symptomatic people. $\xi_c$ is taken as a gamma distribution (18). In the model, the mean and standard deviation are respectively equal to 5 and 2.2 days. |
| Fraction of asymptomatic people | $\alpha_a$ | 0.20 | $\alpha_a$ represents the fraction of asymptomatic with respect to symptomatic patients. $\alpha_a = 0.2$ corresponds to 20% of asymptomatic and 80% of symptomatic. The value taken in this study is given by Mizumoto et al. (19). |
| Recovery time of asymptomatic and symptomatic | $\Delta T_s$ | 14 days | Patients who remained in a given state during the corresponding recovery time is automatically switched to the "removed" state. |
| Recovery time of critical | $\Delta T_c$ | 21 days | |
| Recovery time of severe | $\Delta T_I$ | 14 days | |
| Switching period from symptomatic to severe | $\delta T_s$ | 4-10 days | Time window during which a patient may switch to the next evolution state. For example, a "severe" patient may switch to "critical" from day 0 to day 4. After day 4, the patient is supposed medically stabilised. The switching date is randomly drawn in the time window. |
| Switching period from severe to critical | $\delta T_c$ | 0-4 days | |
| Switching period from critical to deceased | $\delta T_d$ | 2-4 days | |
| Switching probability from symptomatic to severe | $p_s$ | 0.14 | Probability to switch to the newt state during the switching period. |
| Switching probability from severe to critical | $p_c$ | 0.25 | |
| Switching probability from critical to deceased | $p_d$ | 0.35 | |

Without loss of generality, we analyse the simulated number of daily new infected people shown in Figure 6 A. The first half of the curve regularly decreases, indicating that the epidemic spread control is efficient. A conspicuous change of slope occurs at time 35 where a clear change of trend is visible. However, this change of slope is seen with a delay of 5 days since we started to increase *R*_0_ at time 30. In order to test whether our method is able to detect the change of *R*_0_ at time 30, we use a set of control points {*d*_10_*,…,d*_25_} to fit the trend model in the bootstrap procedure (black dots in Fig. 6 A). These control points belong to a period where the situation is stable with a good damping of the epidemic spread. The points to be tested go from day 26 to day 40. The points {*d*_26_*,…,d*_30_} shown as green dots in Fig. 6 A) belong to the stable period like the control points, and the data points {*d*_31_*,…,d*_40_} belong to the beginning of the destabilisation period when *R*_0_ start to increase slowly (red dots in Fig. 6 A).

**Fig. 6.**
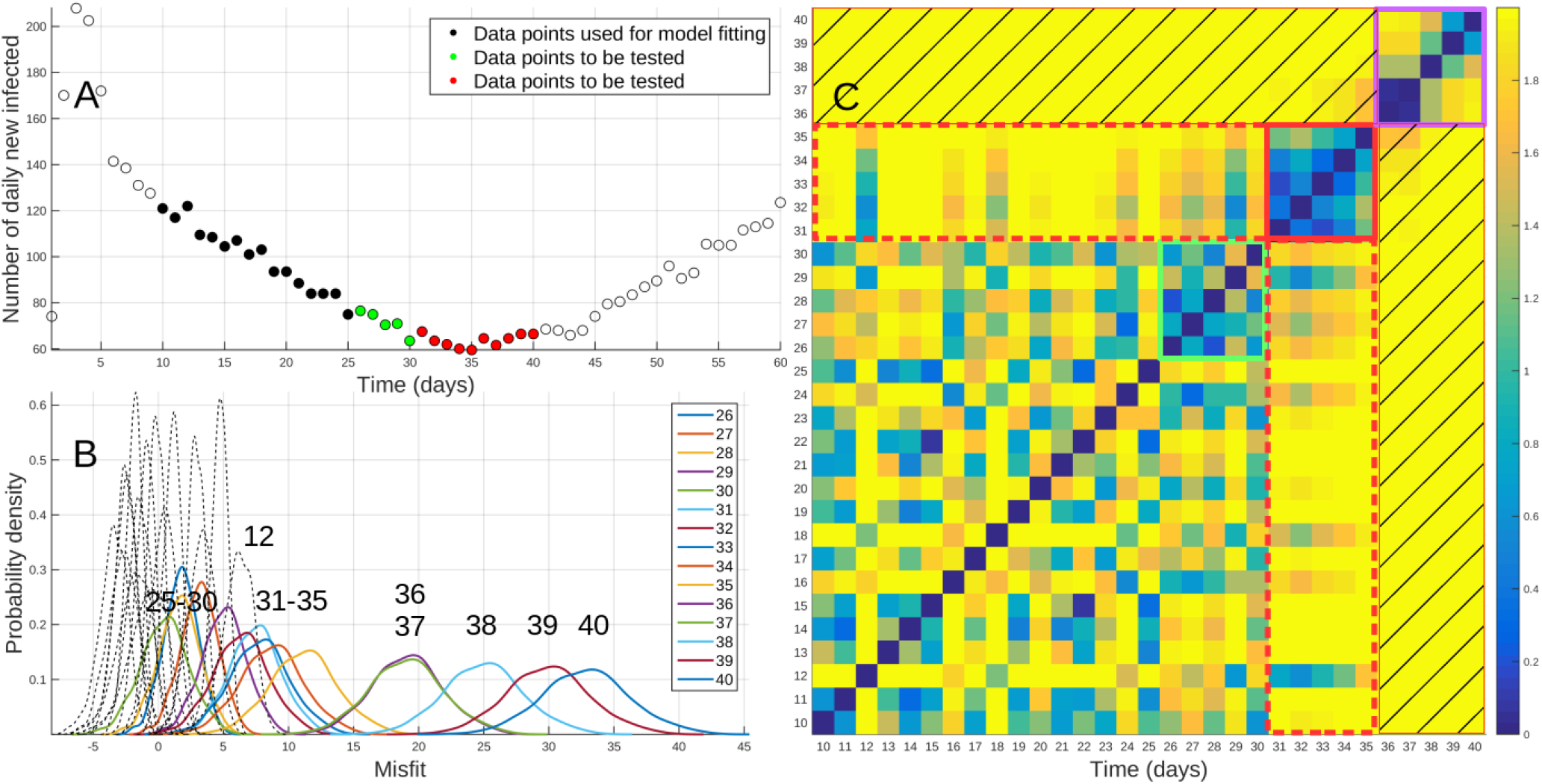
Analysis of a data corresponding to a slow destabilisation of spread control. A) Time series of the daily number of new infected people. Filled black dots = control points used to fit the trend model, filled green and red dots = data points to be tested. B) Probability density functions obtained by bootstrap. Thin dashed curves = pdf’s of data used to fit the trend model, coloured curves = pdf’s of data to be tested. C) Distance matrix **D** (see text for explanations of marked areas). Colour bar range from 0 to 2.

The bootstrapped probability density functions are shown in Figure 6B where the dashed black curves represent the pdf’s of the control points and the coloured curves represent the pdf’s of the test points. The distance matrix is displayed in Figure 6C in its natural order form **D**. In the present example, the rearranged distance matrix **D**\* is not particularly informative and is not shown. The distance matrix contains several interesting features, some of them are emphasised with coloured rectangles in Figure 6C.

The principal features visible in the distance matrix are: i) a large square, with a coherent texture, going from day 10 to day 30; ii) a small square area of coherent texture, enclosed in red, going from day 31 to day 35; iii) a small square area with a more heterogeneous texture, enclosed in violet, going from day 36 to day 40.

The large square going from day 10 to day 30 contains all control data points including the test points {*d*_26_*,…,d*_30_} enclosed by a green square. This indicates that these test points do not significantly depart from the trend constrained by the control points. This is normal since these points belong to the stable period where *R*_0_ = 1.5. As can be checked in Figure 6B, the pdf’s of these test points largely overlap the pdf’s of the control points.

The area enclosed in red square corresponds to the test points {*d*_31_*,…,d*_35_} belonging to the very beginning of the destabilisation period. The distances between these points and the control points are contained in the two red dashed rectangles that are mainly filled with yellow pixels, i.e. corresponding to large distances. This clearly indicates that the set of test points {*d*_31_*,…,d*_35_} significantly departs from the trend defined by the control points. Consequently, we may safely conclude that “something happened” at day 31. The fact that the corresponding pdf’s are located on the right side with respect to the pdf’s of the control points indicates that there is a positive bias in the misfits. Such a positive bias is typical of a decrease of the slope of the trend, indicative of a route to a possible destabilisation.

The area enclosed by a violet square corresponds to the test points {*d*_36_*,…,d*_40_}, i.e. during a period where *R*_0_ continues to increase, making the situation en route toward the exponential divergence after passing the critical boundary at the tipping point which occurs at day 35 with *R*_0_ = 1.625. For these test points, the disagreement with the trend model is even more important, and the distances of these points with all other points are large (i.e. yellow pixels in the hatched rectangles). These test points thus firmly confirm the occurrence of the change announced with the analysis of the test points {*d*_31_*,…,d*_35_}.

A last thing that can be found in the distance matrix of Figure 6C are the presence of outliers. Such is the case of data point *d*_12_ which appears isolated from the other control points in the sense that line 12 and column 12 are principally filled with yellow pixels corresponding to the largest distances. The same is observed to a lesser extent for data point *d*_18_.

A strategy of on-line monitoring is shown in Figure 7 where a sliding procedure is applied to the data. In the top row of the Figure, the procedure starts with given sets of control points {*d*_6_*,…,d*_20_} and test points {*d*_21_*,…,d*_25_} (Fig. 7A1). The corresponding pdf’s and the distance matrix show that both sets of control and test points are mutually coherent and that no clear change point appears (Fig. 7B1-C1). In the next iteration of the sliding procedure, the test points of the preceding iteration are included in the set of control points {*d*_11_*,…,d*_25_} and new test points {*d*_26_*,…,d*_30_} are considered (Fig. 7A2). Again, no clear change point is visible, and we proceed with the next iteration of the procedure where control points {*d*_16_*,…,d*_30_} and new test points {*d*_31_*,…,d*_35_} (Fig. 7A3). Now, the set of test points appears as an isolated self-coherent blue square in the upper-right corner of the distance matrix (Fig. 7C3). This indicates that the test points are incoherent with respect to the control points as previously seen in the discussion of Figure 6.

**Fig. 7.**
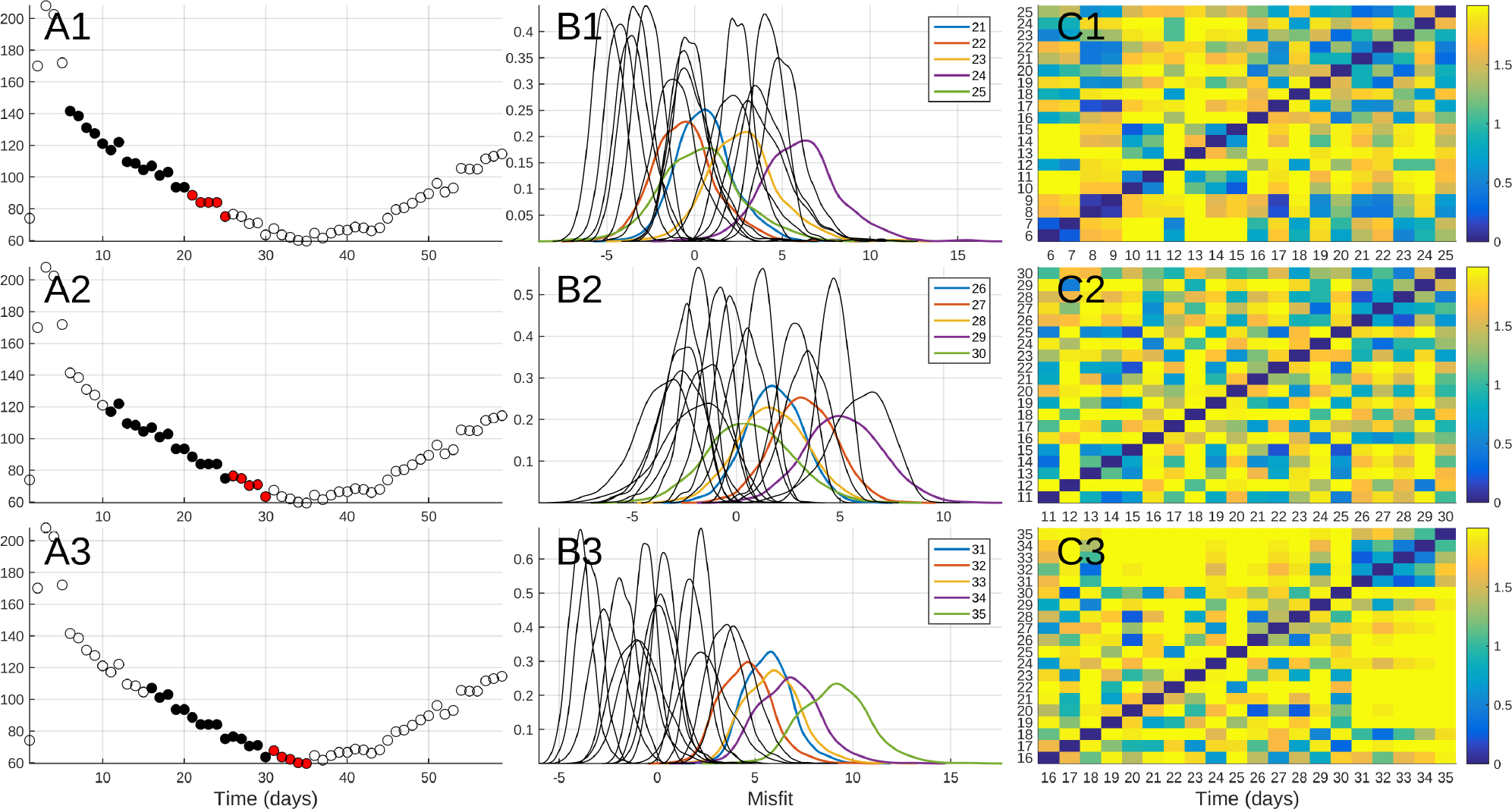
Strategy of on-line monitoring with a sliding procedure where the sets of control and test points are moved along the time axis at each iteration. A1) First iteration with control points {*d*_6_*,…,d*_20_} (filled black dots) and test points {*d*_21_*,…,d*_25_} (red dots). B1) Corresponding pdf’s for a linear trend model. C1) Distance matrix **D** where the set of test points does appears coherent with the control points. A2-C2) Second iteration of the procedure with control points {*d*_11_*,…,d*_25_} and test points {*d*_26_*,…,d*_30_*g*. A3-C3) Third iteration with control points {*d*_16_*,…,d*_30_} and test points {*d*_31_*,…,d*_35_*g*. The set of test points appears as an isolated blue square in the upper-right part of the distance matrix (C3), indicating that a change-point occurred at day 31.

To conclude with this Section, we may claim that our method is able to reliably detect subtle changes in the trend of the time-series of daily new infected persons (Fig. 6A). This detection was possible as early as day 30, i.e. precisely when *R*_0_ begins to increase slowly and five days before a change of trend becomes visible (i.e. at time 35).

## Analysis of a Guadeloupe data set

The data used in the present study are shown in Figure 8A. They correspond to the cumulative number of infected persons with COVID-19 in Guadeloupe and presenting severe symptoms. The time series goes from March 13, 2020 to May 15, 2020. The data are communicated every day by the University Hospital and sanitary services to the Regional Health Agency (Agence Régionale de Santé in French). Data for France are available of the web site of Santé Publique France (15) (see also Alamo et al. (16) for a review of open data repositories).

**Fig. 8.**
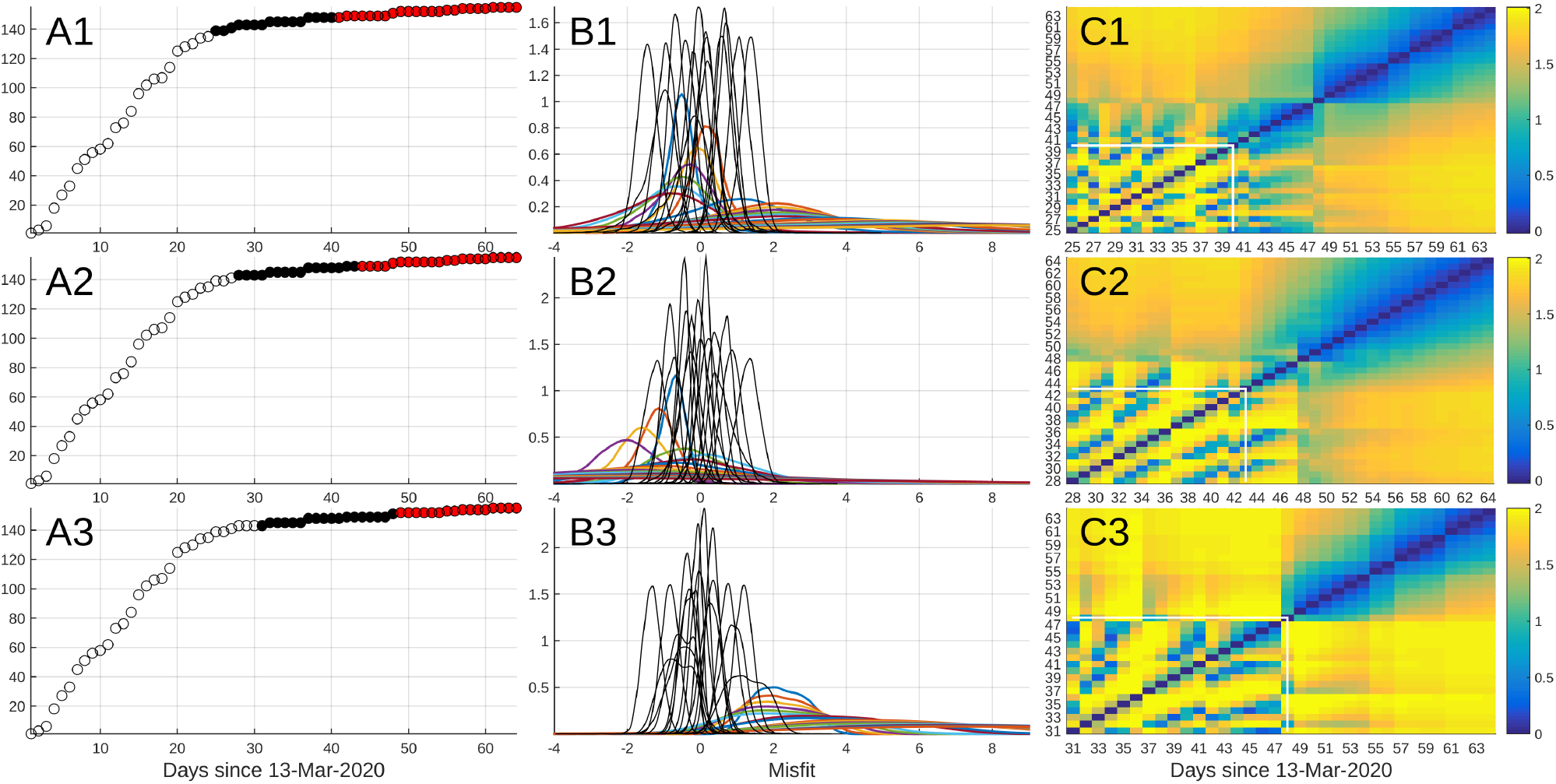
Analysis of the data of Guadeloupe with a sliding procedure as in Figure 7. A1) First iteration with control points {*d*_25_*,…,d*_40_} (filled black dots) and test points {*d*_41_*,…,d*_64_} (red dots). B1) Corresponding pdf’s for a linear trend model. C1) Distance matrix **D** where the control points enclosed by a white rectangle are coherent with the test points {*d*_41_*,…,d*_48_*g*. The test points {*d*_49_*,…,d*_64_} constitute a disjoint blue self-coherent area in the upper-right part of **D**. A2-C2) Second iteration of the procedure with control points {*d*_28_*,…,d*_43_} and test points {*d*_44_*,…,d*_64_*g*. A3-C3) Third iteration with control points {*d*_31_*,…,d*_48_} and test points {*d*_49_*,…,d*_64_}. In all iterations, the test points {*d*_49_*,…,d*_64_} are clustered in an isolated blue area in the upper-right part of the distance matrix, indicating that a change-point occurred near day 49 (i.e. April 30, 2020). Colour bar range from 0 to 2.

Three iterations of a sliding procedure in the same spirit of the one presented in Figure 7 are shown in Figure 8. At each iteration, the set of control points (black dots) is moved along the time axis by a step of 3 days, and the remaining data are all included in the set of test points (red dots). In order to account for the curvature of the time-series formed by the control points, we use a polynomial of degree 2 as trend model. The bootstrapped pdf’s and the distance matrices are shown in Figure 8B and C respectively.

The first iteration is performed with control points {*d*_25_*,…,d*_40_} and test points {*d*_41_*,…,d*_64_} (Fig. 8A1). The pdf’s of the control points are well-grouped (Fig. 8B1) and the distance matrix (Fig. 8C1) shows that the set of test points is divided in two subsets. The first one counts the test points {*d*_41_*,…,d*_48_} which are grouped in a blue domain that may not be considered significantly disjoint from the set of control points. The second subset of test points {*d*_49_*,…,d*_64_} constitutes a disjoint blue self-coherent area in the upper-right part of **D**. In next two iterations, the incoherent characteristic of the data points {*d*_49_*,…,d*_64_} is confirmed. This indicates that a change-point is likely to have occurred near day 49 (i.e. April 30, 2020).

The similarity between the Guadeloupe results (Fig. 8) and the synthetic examples (Fig. 5 and 7) allows to suspect that a subtle change occurred in the data around April 30, 2020. The right-shifted position of the pdf’s after this date (Fig. 8B3) indicate that the change is in the sense of a slight acceleration of the number of infected people. This may be interpreted as a small increase of the basic reproduction number *R*_0_. Owing to the fact that the data are the cumulative number of infected persons presenting severe symptoms (i.e. sick enough to go see the doctor), a delay of about 2 weeks is likely between the change of *R*_0_ and the occurrence of the subtle change detected on April 30 ((5)). This would place the *R*_0_ change near April 15, i.e. approximately at the date when French President Mr Macron announced that the post-containment period will start on May 11, 2020. This coincidence may reflect the fact that the announcement of the containment end made people more confident and less strict in their compliance with the sanitary rules. If we accept this scenario, we find that the basic reproduction number should be augmented to *R*_0_ ≈ 0.7 after April 13–15.

## Concluding remarks

The method proposed in the present study is easy to implement and relies on simple statistical considerations. The computing time is very short and the results are displayed in an easily understandable form (e.g. Fig. 5).

The simulation test of Figure 6 shows that early warning signals announcing small changes in the conditions of the epidemic spread control may be detected several days before the appearance of a visually clear change point (Fig. 6A). Such a possibility may help to monitor the evolution of the epidemic spread during the post-containment period where it is of a primary importance to detect any disturbance of the spread control as soon as possible.

The analysis of the Guadeloupe data allows to suspect that a subtle disturbance occurred in the data around April 30, 2020. The most recent data available at the time of completion of the present article show that the disturbance of April 30 is not due to outliers but instead seems attributable to a persistent change in the trend. This may be due to an increase of *R*_0_ ≈ 0.7 around April 13–15, 2020.

## Data Availability

The data are available on the website of Sante Publique France

## ACKNOWLEDGEMENTS

The MatLab^®^codes are available upon request. No dedicated funds were attributed to this study.

## Supplementary Note 1: Model parameters

